# A novel deep learning method for large-scale analysis of bone marrow adiposity using UK Biobank Dixon MRI data

**DOI:** 10.1101/2022.12.06.22283151

**Authors:** David M Morris, Chengjia Wang, Giorgos Papanastasiou, Calum D Gray, Sammy Badr, Julien Paccou, Scott IK Semple, Tom MacGillivray, William P Cawthorn

## Abstract

**OBJECTIVES:** Bone marrow adipose tissue (BMAT) represents >10% of total fat mass in healthy humans and further increases in diverse clinical conditions, but the impact of BMAT on human health and disease remains poorly understood. Magnetic resonance imaging (MRI) allows non-invasive measurement of the bone marrow fat fraction (BMFF), and human MRI studies have begun identifying associations between BMFF and skeletal or metabolic diseases. However, such studies have so far been limited to smaller cohorts: analysis of BMFF on a larger, population scale therefore has huge potential to reveal fundamental new knowledge of BMAT’s formation and pathophysiological functions. The UK Biobank (UKBB) is undertaking whole-body MRI of 100,000 participants, providing the ideal opportunity for such advances.

**MATERIALS AND METHODS:** Herein, we developed a deep learning pipeline for high-throughput BMFF analysis of these UKBB MRI data. Automatic bone marrow segmentation was achieved by designing new lightweight attention-based 3D U-Net convolutional neural networks that allowed more-accurate segmentation of small structures from large volumetric data. Using manual segmentations from 61-64 subjects, the models were trained against four bone marrow regions of interest: the spine, femoral head, total hip and femoral diaphysis. Models were validated using a further 10-12 datasets for each region and then used to segment datasets from a further 729 UKBB participants. BMFF was then determined and assessed for expected and new pathophysiological characteristics.

**RESULTS:** Dice scores confirmed the accuracy of the models, which matched or exceeded that for conventional U-Net models. The BMFF measurements from the 729-subject cohort confirmed previously reported relationships between BMFF and age, sex and bone mineral density, while also identifying new site- and sex-specific BMFF characteristics.

**CONCLUSIONS:** We have established a new deep learning method for accurate segmentation of small structures from large volumetric data. This method works well for accurate, large-scale BMFF analysis from UKBB MRI data and has the potential to reveal novel clinical insights. The application of our method across the full UKBB imaging cohort will therefore allow identification of the genetic and pathophysiological factors associated with altered BMAT. Together, our findings establish the utility of deep learning for population-level BMFF analysis and promise to help elucidate the full impact of BMAT on human health and disease.

**Highlights:**

- We establish a new deep learning method for image segmentation.
- Our method improves segmentation of small structures from large volumetric data.
- Using our method, we assess bone marrow fat fraction (BMFF) in UK Biobank MRI data.
- This is the first use of deep learning for large-scale, multi-site BMFF analysis.
- Our results highlight the potential of BMFF as a new clinical biomarker.

## 1. INTRODUCTION

Bone marrow adipose tissue (BMAT) accounts for up to 70% of total bone marrow (BM) volume and approximately 10% of total fat mass in lean, healthy humans (1). BMAT further increases with ageing and in diverse clinical conditions, including osteoporosis, obesity, type 2 diabetes, oestrogen deficiency, chronic kidney disease, radiotherapy and glucocorticoid treatment (1). In striking contrast to other adipose depots, BMAT also increases during caloric restriction in animals and in humans with anorexia nervosa (1-4). Thus, BMAT is a major component of normal human anatomy; is distinct to other types of adipose tissue; and is altered in numerous clinical contexts.

These observations suggest roles for BMAT in normal physiological function and the pathogenesis of multi-morbidities, including major ageing-associated diseases. Indeed, clinical and preclinical studies suggest that BMAT can directly influence skeletal remodelling, haematopoiesis and energy homeostasis (1, 5, 6) and have revealed endocrine properties through which BMAT may exert systemic effects (3). However, study of BMAT has been limited, especially in comparison to other major adipose depots (1); hence, BMAT formation and function remains poorly understood. Despite this relative ignorance, recent studies have revealed new fundamental knowledge of BMAT biology. One key finding is that BMAT’s characteristics and functions differ according to its skeletal location. BMAT is proposed to exist in two broad subtypes, dubbed ‘constitutive’ and ‘regulated’ (7, 8): constitutive BMAT predominates in the appendicular skeleton, particularly at more-distal sites, whereas regulated BMAT develops in the axial skeleton and in proximal regions of the long bones, such as the femoral head and epiphysis. Adipocytes within regulated BMAT increase or decrease in size and/or number in response to altered environmental, physiological and pathological conditions, whereas those within constitutive BMAT are relatively resistant to expansion or breakdown in such contexts (7, 8). Thus, efforts to further elucidate BMAT formation and function must consider these fundamental site-specific differences.

Magnetic resonance imaging (MRI) and proton MR spectroscopy have emerged as key tools for non-invasively assessing BMAT properties in humans (9), including the extent of BM adiposity and the proportions of saturated and unsaturated lipids within the BM (10). The former depends on analysis of BM fat fraction (BMFF) using chemical shift-encoding based water-fat separation methods. These approaches have been applied in various small- and mid-scale human cohort studies, revealing some insights into BMAT’s association with human skeletal and metabolic health (11, 12). For example, multiple studies have shown that BMFF is increased in osteoporosis and is associated with lower bone mineral density (BMD) in non-osteoporotic subjects (11-13). However, these cohort studies have never included more than 560 people (13), limiting the ability to detect other associations. Thus, analysis of BMFF on a larger scale has enormous potential to reveal fundamental new knowledge of BMAT formation and function, including the association with other physiological, pathological and genetic variables. This would provide new understanding about the factors that regulate BMAT development, as well as highlighting how altered BMFF impacts human health and disease.

The UK Biobank (UKBB) is undertaking the world’s largest health imaging study (14), providing an ideal opportunity for such large-scale BMFF analysis. Of the 500,000 UKBB participants, 100,000 are undergoing MRI of the brain, heart and whole body, as well as dual-energy X-ray absorptiometry to measure BMD. As of August 2022, approximately 53,000 participants have been scanned. Efficient measurement of BMFF from these MRI datasets will require development of new automated analysis methods. Several groups have developed machine learning for automated segmentation of other anatomical regions from the UKBB MRI data (15-17). Machine learning has also recently been used to segment the knee or vertebral BM from Dixon images in smaller cohorts outwith the UKBB (18-20); however, machine learning has not yet been developed for automated segmentation of the BM from other skeletal sites, and never using MR data from the UKBB. These were the goals of the present study.

Given the potential insights that could be gained from such large-scale BMFF analysis, herein we developed a deep learning pipeline for automated BM segmentation from UKBB MRI data. Our findings establish the utility of deep learning for large-scale analysis of BMFF within the UKBB and the potential of this approach for revealing the impact of BMAT on human health and disease.

## 2. MATERIALS AND METHODS

### 2.1. UKBB Imaging study – participants

Full details of the UKBB imaging study have recently been reported by Littlejohns *et al*, who summarise the study as “*a population-based cohort of half a million participants aged 40–69 years recruited between 2006 and 2010. In 2014, UK Biobank started the world’s largest multi-modal imaging study, with the aim of re-inviting 100,000 participants to undergo brain, cardiac and abdominal magnetic resonance imaging, dual-energy X-ray absorptiometry and carotid ultrasound*” (14). As of August 2022, over 53,000 participants have undergone the UKBB imaging protocol. The phenotypic and imaging data used in this study were obtained from UKBB and analysed under an approved project application (ID 48697). All work reported herein was done in accordance with UKBB ethical requirements.

### 2.2. UKBB – MRI acquisition

MRI data were acquired on a 1.5 T whole-body MR system (Magnetom Aera, Siemens Medical Solutions, Erlangen, Germany). Tridimensional two-point Dixon sequences were used to give coverage from neck to knees, consisting of six volumes. In the present study we analysed three of these volumes: the abdomen, hips, and upper leg. For the abdomen and hips, breath-hold sequences were acquired by using a 3D dual-echo spoiled gradient-echo (FLASH) T1-weighted acquisition using the following parameters: TR/TE_in-phase_/TE_out-of-phase_: 6.7/4.8/2.4 ms; field of view (FOV): 500 × 381 mm; slice thickness: 4.5 mm; isotropic in-plane spatial resolution of 2.2 mm; number of slices: 44. Parallel imaging factor 2 in both frequency/phase directions and a partial Fourier reconstruction of 71% were used to reduce acquisition time. For the upper leg slice, slice thickness was reduced to 3.5 mm and 72 slices were acquired with the same resolution. Detailed technical parameters are available in the UKBB rationale (14).

### 2.3. UKBB – DXA scans for bone mineral density measurement and body composition

As part of the UKBB Imaging study, bone mineral density (BMD) was measured at the lumbar spine (L1–L4) and at the non-dominant hip for femoral neck and total hip by DXA scan (GE-Lunar iDXA). Machines were calibrated daily, and quality-assurance tests were carried out periodically. WHO criteria were used to define osteoporosis (BMD T-score ≤−2.5) and osteopenia (BMD T-score between −1.0 and −2.5). All UKBB imaging participants also underwent total-body DXA scanning (GE-Lunar iDXA). Fat, lean, and bone masses for the total body and per region (arms, legs, and trunk) were measured and analyzed using the manufacturer’s validated software, with visceral adipose tissue (VAT, kg) also measured. Daily quality-control and calibration procedures were performed using the manufacturer’s standards.

### 2.4. Training and validation cohort

To develop a deep learning method for automated BM segmentation we focussed on a subset of UKBB Imaging participants, consisting of 729 male and female subjects aged 60-69 years old (Table 1). This cohort was selected to include control subjects (with normal BMD) and subjects with osteopaenia or osteoporosis. Subjects with obesity and type 2 diabetes were excluded because these conditions can influence BMFF (1, 6), leaving only non-diabetic subjects with a body mass index (BMI) within the normal range (18.5-25 kg/m^2^).

**Table 1.** Characteristics of subjects in training and validation cohort. Normally distributed data are reported as mean ± SEM while non-normally distributed data are reported as median [interquartile range]. BMI, body mass index; DXA, dual-energy X-ray absorptiometry; VAT, visceral adipose tissue. Within each sex, significant differences between control subjects and osteopaenic or osteoporotic subjects are indicated by * (*P* <0.05), ** (*P* <0.01) or *** (*P*<0.001). Within control subjects, significant differences between males and females are indicated by ^##^ (*P* <0.01) or ^###^ (*P* <0.001).

|  | **Males (n = 277)** |  |  | **F (n = 452)** |  |  |
| --- | --- | --- | --- | --- | --- | --- |
|  | **Control**<br>**(n=138)** | **Osteopaenic**<br>**(n=146)** | **Osteoporotic**<br>**(n=17)** | **Control**<br>**(n=134)** | **Osteopaenic**<br>**(n=262)** | **Osteoporotic**<br>**(n=70)** |
| Age (years) | 65 [63, 67] | 65 [63, 67] | 64.47 ± 0.7 | 65 [62, 67] | 65 [62, 67] | 65 [63, 67] |
| BMI (kg/m<sup>2</sup>) | 23.6 [22.8, 24.3] | 23.3 [22.3, 24.1] | 22.04 ± 0.20 *** | 22.9 [21.7, 23.9] ## | 22.6 [21.3, 23.7] | 21.67 ± 0.40 * |
| BMD T-score (L1-L4) | 0.65 [-0.2, 1.775] | -1 [-1.575, -0.1] *** | -3 [-3.25, -1.55] *** | 0.15 [-0.4, 0.9] | -1.5 [-1.9, -0.8] *** | -2.8 [-3.1, -2.6] *** |
| BMD T-score (total femur, left) | 0.2 [-0.3, 0.7] | -1.12 ± 0.05 *** | -2.2 ± 0.14 *** | 0 [-0.4, 0.475] | -1.4 [-1.8, -1] *** | -2.22 ± 0.09 *** |
| BMD T-score (femoral neck, left) | -0.3 [-0.7, 0.275] | -1.5 [-1.8, -1.2] *** | -2.45 ± 0.13 *** | -0.15 [-0.7, 0.4] | -1.45 [-1.8, -1.1] *** | -2.11 ± 0.07 *** |
| Android tissue fat % by DXA | 30.6 [24, 34.6] | 30.0 [22.8, 35.7] | 24.4 ± 2.0 | 34.8 [27.8, 40.7] ### | 32.5 ± 0.6 | 31.0 ± 1.1 |
| Gynoid tissue fat % by DXA | 24.3 ± 0.4 | 24.4 ± 0.4 | 23.5 ± 1.0 | 37.6 ± 0.4 ### | 38.5 ± 0.3 | 38.7 ± 0.6 |
| Legs tissue fat % by DXA | 20.9 ± 0.3 | 21.2 ± 0.3 | 21.3 ± 1.0 | 35.2 ± 0.5 ### | 36.9 ± 0.3 | 37.1 ± 0.6 |
| Trunk tissue fat % by DXA | 29.1 [23.7, 32.0] | 28.6 [23.0, 33.4] | 24.3 ± 1.5 | 35.4 [29.9, 39.5] ### | 33.3 ± 0.4 | 32.3 ± 0.9 |
| Total tissue fat % by DXA | 24.6 ± 0.4 | 25.6 [21.6, 28.5] | 22.9 ± 1.8 | 34.7 [30.9, 37.38] ### | 34.3 ± 0.3 | 33.9 ± 0.6 |
| VAT mass (g) | 949.4 ± 35.25 | 783.5 [465.5, 1131] | 586 ± 79.6 ** | 407 [225.5, 717] ### | 346.5 [217, 563.5] | 296 [193.3, 526.5] |

### 2.5. Data management and workflow

MRI data was downloaded from UKBB, consisting of multiple volumes acquired using the 2-point Dixon technique, based on the parameters listed above. For each volume the in- and out-of-phase, fat and water images were available. The data were downloaded in flat format and sorted by sequence to expedite data access. The volumes required were identified by their sequence number assuming a standard acquisition protocol, which was determined from the data. As shown in Figure 1, we began by downloading and analysing data from the 729-subject training and validation cohort.

**Figure 1.**
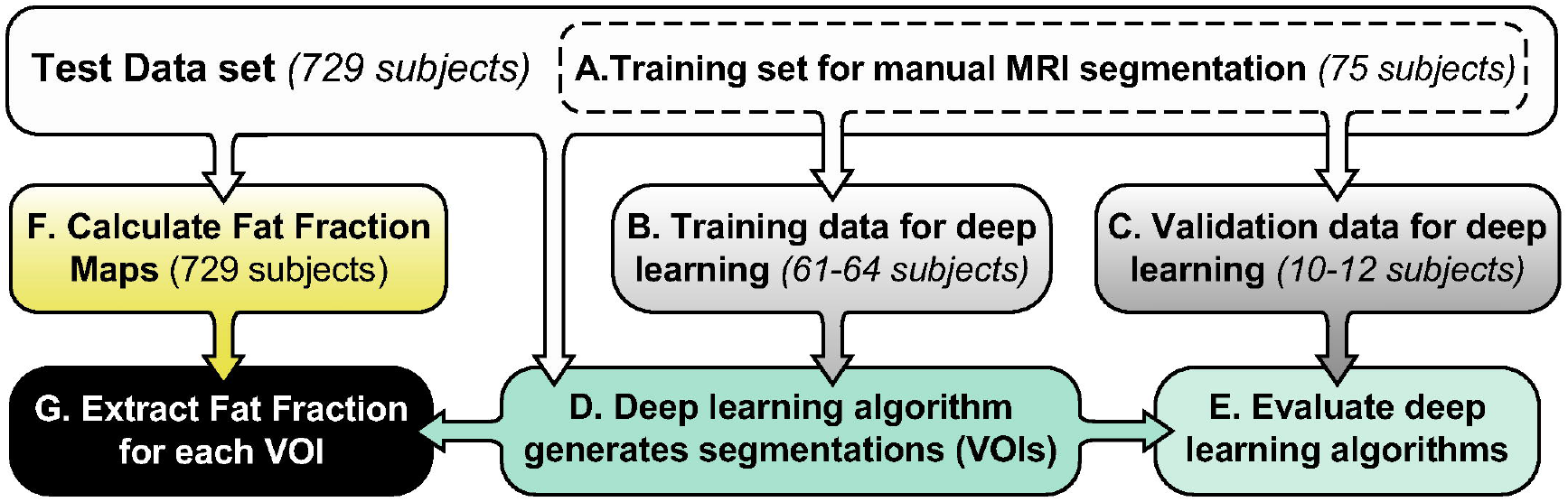
Workflow for data management, manual segmentation and application and validation of deep learning. The test dataset comprised the validation cohort of 729 subjects (described in Table 1), from which datasets from 75 subjects were manually segmented (A) to generate four VOIs per subject (spine, femoral head, total hip, and femoral diaphysis). The manual segmentations from 61-64 of these subjects were used to train the deep learning models for each VOI (B), while those from 10-12 subjects were kept as ‘unseen’ segmentations that had not been used to train the models (C). The models were then used to segment all datasets from the 729-subject cohort (D), with deep learning segmentations from the 10-152validation datasets then compared to the corresponding manual segmentations to calculate dice coefficients for each model (E). Finally, FF maps were generated from each MRI dataset (F) and the deep learning segmentations applied to these to obtain the BMFF for each VOI (G).

### 2.6. Manual segmentation of MRI data

A training dataset of 75 subjects (Fig. 1A) was extracted from the test dataset to be used for the training and validation of the deep learning algorithms. Each of these 75 datasets was segmented by a single observer for consistency, generating manual segmentations. For each subject, the fat images were used to define four distinct volumes of interest (VOIs) corresponding to BM regions of pathophysiological relevance: the spine, the femoral head, the total hip, and the femoral diaphysis. The spine consisted of all the vertebral marrow in the principle abdominal volume, which contained 6-7 vertebrae ranging from T8 to L3. The reason for this range of vertebrae is that the multiple abdominal acquisitions have a fixed volume and are continuous across the patient’s body; hence, the range of vertebrae within each abdominal volume depends on the patient’s height. The femoral head and total hip regions were segmented from the hip volume. Here, the total hip consisted of the femoral neck and the hip between the lesser and greater trochanter. The femoral diaphysis, located in the upper leg volume, was segmented at the mid-shaft of the femur, which was identified by locating the point of the shaft with the narrowest cross section. Each femoral volume was segmented from the non-dominant left femur to allow more-direct comparison with DXA measurements, which are usually performed at the non-dominant hip. Femoral BMFF does not show significant contralateral differences (21), meaning that BMFF measurements from the left femur should be representative of both sides. Segmentation was performed on the native axial images on a slice-by-slice basis in Analyze 12.0 software (AnalyzeDirect, Overland Park, KS, USA) following an overall inspection of each volume to determine the extent of each region excluding partial volume, defined as a drop in signal intensity > 50% compared to the centre of the region.

Of the 75 manually segmented datasets (Fig. 1A), 64 were used to train the deep learning model for the spine; 61 were used for the femoral head and diaphysis; and 62 were used for the total hip (Fig. 1B). To do so, the fat images and their corresponding manual segmentations were used iteratively to build a separate model to segment each region individually and generate a deep learning segmentation (Fig. 1D). The remaining datasets (Fig. 1C) were not used in training the models but instead were used as unseen validation data to test the models: 12 datasets were used for testing the spine, 11 for the femoral head, and 10 each for the total hip and diaphysis models. For these validation datasets, comparison of the deep learning segmentations with the manual segmentations (Fig. 1E) allowed dice coefficients to be calculated for the four different algorithms (Table 2).

**Table 2.** Segmentation Accuracy (dice scores) of the traditional U-Net and our CBAM-ROI-attention U-Net.

|  | **Vertebrae** | **Femoral head** | **Total Hip** | **Femoral Diaphysis** |
| --- | --- | --- | --- | --- |
| **U-Net** | 0.925 | 0.951 | 0.904 | 0.69 |
| **ROI-Attention-U-Net** | 0.912 | 0.945 | 0.912 | 0.866 |

All the deep learning segmentations for the training and validation datasets were manually checked. This identified several data issues and segmentation failures that required the development of specific error-checking rules. These rules were based on determining if the VOIs generated were physiologically appropriate: VOIs could not consist only of single voxels, nor were gaps allowed within the VOIs. Therefore, the initial error-checking steps automatically removed any single-voxel VOIs and joined together any discontinuous VOIs. Additional error checking was used to identify those segmentations that were outliers within the distribution of regions generated. This was based on the centre of mass being greater than 3 standard deviations from the mean of the training dataset. This is useful for identifying erroneous segmentations that have been caused by data quality issues or deviations from the standard MRI protocol.

### 2.7. U-Net design and rationale

Directly segmenting 3D data using a traditional U-Net (22) has several drawbacks: i), the size of input data and the depth of the model are limited by the available GPU memory; ii), due to the highly imbalanced distribution between classes, the traditional 3D U-Net (22) tends to label all voxels as background; and iii), the fixed size of the receptive field limited the ability of the model to effectively utilize the global correlations between local features.

To address these issues, we developed a novel light-weight attention-based U-Net model for simultaneous detection and segmentation of tiny structures in large 3D data. Figure 2 shows the architecture of this new Attention ROI U-Net model. The encoding subnetwork output feature maps four resolution levels (23). Each encoding block consists of a conventional U-Net convolutional layer (3D conv + Relu + Instance normalization), a convolutional layer equipped with a modified convolutional block attention module (CBAM) (24), and a down-sampling layer implemented as a stride 2 3×3×3 convolution operation. The last encoding block consists of two CBAM convolutional layers with a non-local spatial attention layer (25) inserted between them. Unlike the original CBAM, which generates two attention maps using average and max pooling, we used 1×1×1 convolution to generate one single fixed-size attention map from each CBAM layer. The 5 attention maps are all resized to 96×96×96 and then fused by a mini convolutional neural network (CNN) with a *Softmax* layer to generate a probability map ***P***. The centre, (*x*, *y*, *z*)_*ROI*_, of a region of interest (ROI), which indicates the location of the segmented anatomical structure, is then given by:

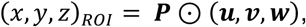

Here, ***u***, ***v***, ***w*** are grid of data coordinates normalized to [-1, 1]. With this centre, a cubic ROI is extracted from the encoder feature maps of all resolution levels with sizes 32, 16, 8 and 4. The U-Net decoder then generates the segmentation of this ROI. The final segmentation results are produced by recovering the ROI location within the original data volume.

**Figure 2.**
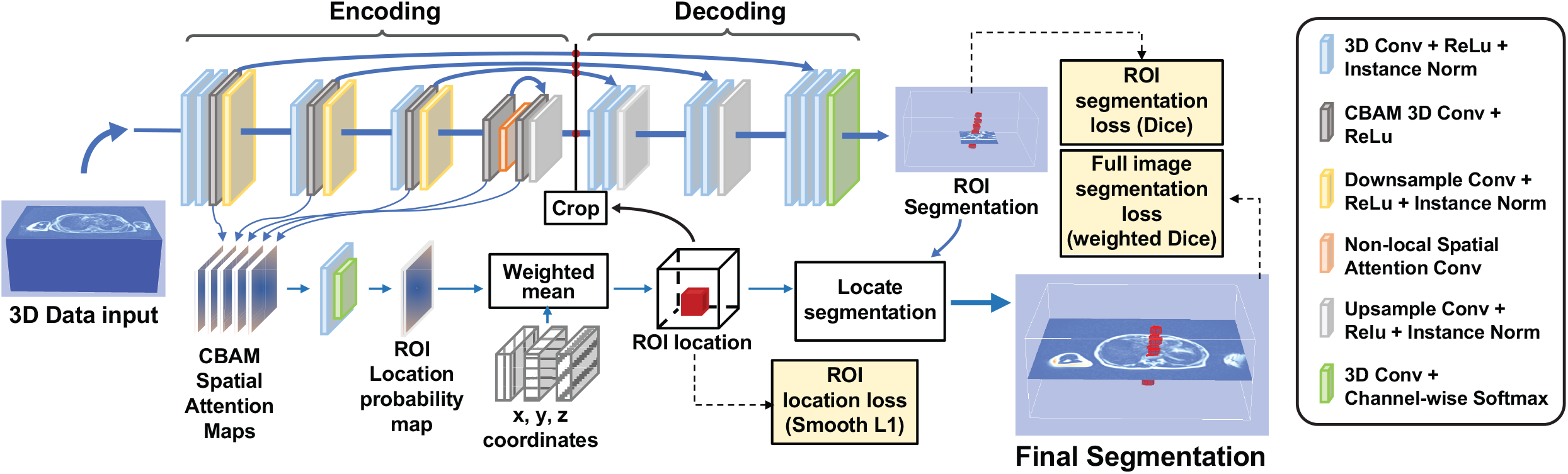
Architecture of our CBAM Attention ROI U-Net for segmenting small structures from large 3D data. Each convolutional block in the U-Net encoding subnetwork (or contracting path) includes one or two CBAM (convolutional block attention module) layers. A fixed-size single channel spatial attention map is generated by each CBAM layer through 1×1×1 convolutions and trilinear interpolation. These attention maps are then combined to produce a probability map of object location with which a ROI is defined. The encoded features of all resolution-levels are then cropped to the ROI and input into the decoder which produces the segmentation results within the detected ROI. A non-local spatial attention layer is inserted in the final block to generate globally sensitive features. The final segmentation results are then generated by implanting the ROI back into the whole data volume.

Detection of the ROI location is realised by minimizing a ROI centre localization loss, *L*_*loc*_, defined on the predicted and ground-truth ROI centres. We use the conventional Dice loss, *L*_*ROI*_, to optimize the segmentation of the detected ROI. Because minimize bias in traduced by the class imbalance on the final segmentation results, we also compute a weighted Dice loss, *L*_*S*eg_ using the full image segmentation, where the weight of each class is defined as the reciprocal of the number of voxels. To sum up, the loss function for trains ing our new U-Net model is defined as:

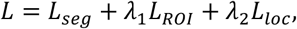

where λ_1_ and λ_2_ control the weights between different losses. In this work, we set *λ*_1_ = *λ*_2_ = 1. The proposed algorithm was implemented in Pytorch (26) with an Adam optimizer (27).

### 2.8. Fat fraction mapping

Fat fraction (FF) measurements from MRI data allow for the determination of the relative quantities of water and fat present within tissue, based on the different resonant frequencies of hydrogen atoms bound to fat and water. Acquisition of in- and out-of-phase images allows fat and water images to be generated. Based on the intensities of these images the FF was calculated as a percent of the voxel volume. This was done for all volumes of interest. The specific VOIs, segmented using our novel U-Net model, were then applied to the FF maps to allow extraction of the FF for each VOI. This used the fat and water images for each volume of interest and nearest-neighbour smoothing was applied to the images before the maps were calculated to minimise the influence of any noise spikes in the data. In house code (Matlab 2019B, The Mathworks Inc, Natick, Massachusetts, USA) applied the deep learning segmentations to the FF maps after erosion of the spine, head and total hip regions by a single boundary voxel in plane to ensure measurements were from marrow and not bone. This erosion step was not applied to the diaphysis segmentations because of the small cross section of this region (for some patients the diaphyseal cross section is so small that it would be eliminated by the erosion step).

### 2.9. Data presentation and statistical analysis

Data were analysed for normal distribution using the Anderson-Darling test. For results tables of summary statistics, normally distributed data are reported as mean ± SEM and were compared using one-way or two-way ANOVA with Šídák’s test for multiple comparisons. Non-normally distributed data are reported as median [interquartile range] and were compared using the Kruskal-Wallis test, with Dunn’s test for multiple comparisons; the latter was also used when comparing normally distributed data with non-normally distributed data. Images of manual and deep learning segmentations were generated using 3DSlicer (v4.11) and colours adjusted using GIMP2. Graphs of summary data are presented as Violin plots overlaid with individual data points. Visualisation and statistical analysis of these summary data were done using Prism software (v9.4.1, GraphPad, USA). Univariable regression analyses were done in RStudio v2022.02.1 (Build 461), with multivariable regression performed using finalfit (R package v1.0.5) (28). Subjects with any erroneous measurements (e.g. a BMD of 0 g/cm^2^) were excluded from the regression analyses. A Bonferroni-adjusted *P*-value <0.05 was considered statistically significant.

### 2.10. Data and code availability

All data for FF and segmentation volumes will be uploaded to the UKBB. Code for the deep learning models will be made available via GitHub. Code for regression analyses will be made available via DataShare (https://datashare.ed.ac.uk). Until these data are publicly available, the authors will agree to all reasonable requests for code and data sharing, in accordance with UKBB guidelines.

## 3. RESULTS

### 3.1. U-Net development and training

We first used MRI data from 61-64 subjects for manual segmentation of four VOIs: the spine, consisting of lumbar and thoracic vertebrae; the femoral head; total hip; and femoral diaphysis. We then trained separate U-Net models for each VOI and tested their performance on 10-12 subjects in a validation dataset (Fig. 1). Figure 2 shows the architecture of our new U-Net, while Table 2 shows the comparison Dice index results between the conventional U-Net and our new U-Net models for each site. Visual comparison of manual vs deep learning segmentations further confirmed the accuracy of the outputs from each of our deep learning models (Fig. 3).

**Figure 3.**
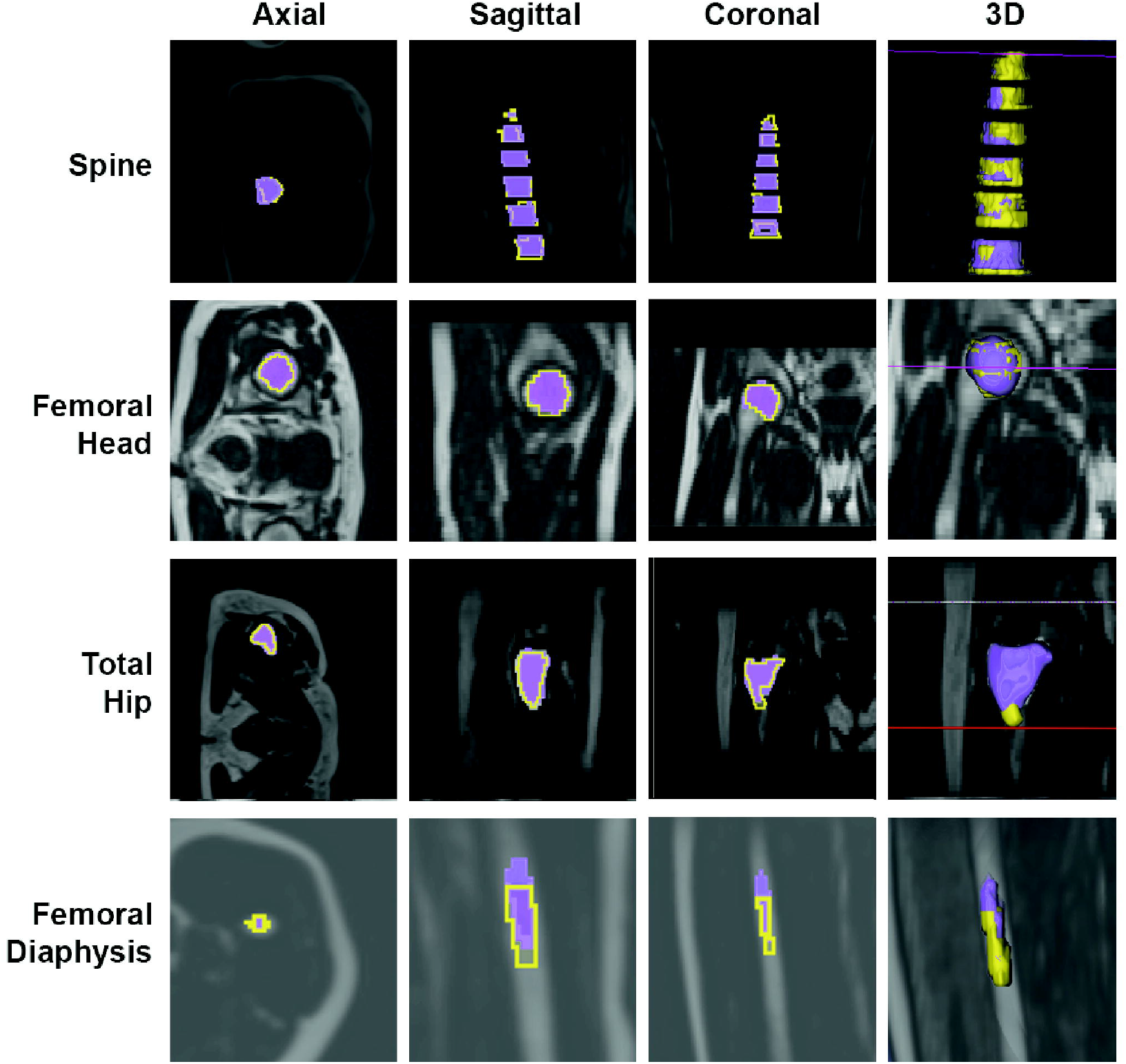
Visual comparison of manual vs deep learning segmentations. Deep learning segmentation results (purple) are displayed on top of the ground-truth (manual) segmentations (yellow). Representative images from the axial, coronal and sagittal plane are shown, along with a 3D rendering.

### 3.2. Segmentation and Fat Fraction mapping of training and validation cohort

To test if our U-Net models yield reliable BMFF results, we next applied them to FF maps from a cohort of 729 UKBB participants (Table 1). This cohort was chosen to include both males and females aged 60-69, comprising individuals with osteoporosis, osteopaenia, or normal BMD. The rationale for this is as follows: first, BMFF increases with age and, for humans aged 60-69, vertebral BMFF is expected to be greater in females than in males (29, 30); second, BMFF is increased in osteoporosis and negatively associated with BMD (1, 6, 12); and finally, BMFF is greater in the femur than in the lumbar spine (1, 31). Thus, applying our U-Net models to analyse spinal and femoral BMFF in this cohort allowed us to test if the resulting deep learning segmentations yield BMFF values that show these expected associations with sex, age, BMD, and anatomical site. If so, this would validate the accuracy of our new models for high-throughput BM segmentation and BMFF analysis.

As shown in Figure 4, we found that BMFF in healthy control subjects significantly differed across the five regions analysed. This was most obvious for the spine, where BMFF was lower than in each femoral region. However, BMFF also differed between each femoral region, being highest in the femoral head and then decreasing progressively in the total hip (*P* = 0.0012 vs femoral head) and diaphysis (*P* <0.0001 vs femoral head or total hip). There were also significant, region-dependent sex differences: spinal BMFF was greater in females than in males, whereas males had greater BMFF at each femoral site (Fig. 4).

**Figure 4.**
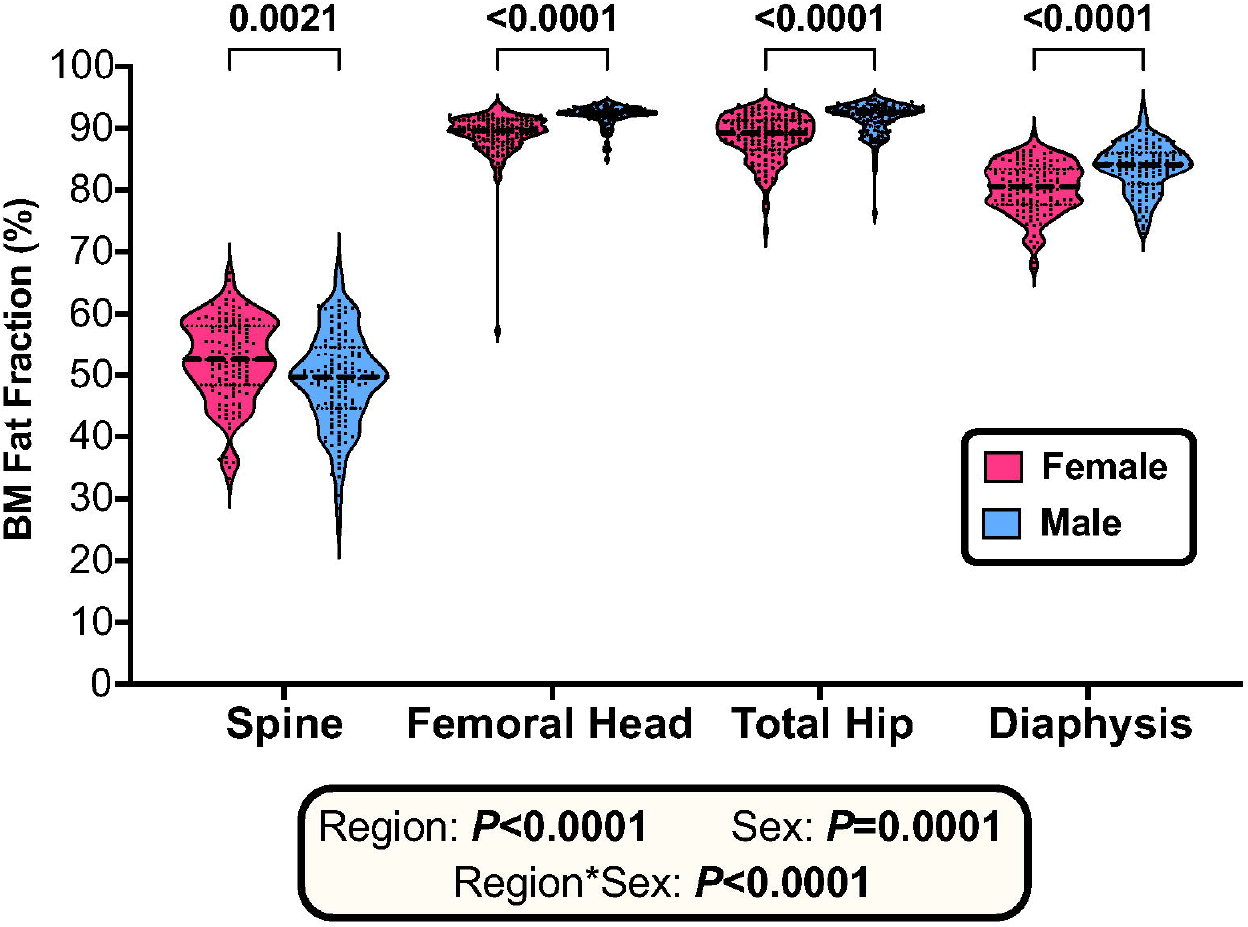
Sex differences in BMFF vary according to skeletal region. BMFF for control subjects was assessed at each skeletal region. Data represent 134 females and 138 males and are shown as violin plots overlaid with individual data points. Significant effects of region, sex, and region*sex interaction were assessed using a mixed-effects model with Šídák’s multiple comparisons test. Overall *P* values for each variable, and their interaction, are shown in the box, while significant sex differences within each region are indicated above.

To further understand the regional and sex differences in BMFF, we investigated if BMFF at one site is associated with BMFF at the other sites. As shown in Table 3, there were strong positive associations between BMFF at each femoral site, with the relationship between total hip BMFF and diaphyseal BMFF being stronger in males than in females. Spinal BMFF was not associated with diaphyseal BMFF; however, it was positively associated with femoral head BMFF in females, and with total hip BMFF in males and females; the latter relationship was also stronger in females than in males (Table 3). Thus, BMFF at one site is generally positively associated with BMFF at other sites, and this relationship differs between the sexes.

**Table 3.** Univariable and sex-stratified associations between BMFF for each region. To test if the explanatory-dependent relationship differs between males and F, a linear model was first analysed across both sexes, with sex included as an interacting variable. Beta coefficients are shown (with lower and upper 95% Cis in brackets), followed by the adjusted R^2^ (Adj. R^2^) and unadjusted *P* value for each explanatory variable (*P* Exp). *P* values were also calculated for the Explanatory*Sex interaction (*P* Exp*Sex); if significant, additional linear models were analysed in females and males separately. Because 12 correlations were assessed, the Bonferroni-adjusted alpha level for *P* (Exp) is 0.05/12 = 0.0042. Significant explanatory-dependent relationships are highlighted in bold.

| Explanatory | Dependent | Sex | $\beta$ (CIs) | Adj. R<sup>2</sup> | P (Exp) | P (Exp*Sex) |
| --- | --- | --- | --- | --- | --- | --- |
| BMFF Spine | BMFF Femoral Head | Both | **0.037 (0.015, 0.059)** | **0.015** | **1.25E-03** | **2.6E-04** |
|  |  | Female | **0.109 (0.08, 0.138)** | **0.118** | **8.69E-13** | - |
|  |  | Male | 0.028 (0, 0.057) | 0.013 | 0.049 | - |
|  | BMFF Total Hip | Both | **0.091 (0.063, 0.12)** | **0.055** | **4.48E-10** | **0.026** |
|  |  | Female | **0.171 (0.132, 0.21)** | **0.145** | **2.16E-16** | - |
|  |  | Male | **0.106 (0.069, 0.144)** | **0.107** | **7.22E-08** | - |
|  | BMFF Diaphysis | Both | 0.054 (0.01, 0.099) | 0.007 | 0.017 | 0.801 |
| BMFF Femoral Head | BMFF Total Hip | Both | **1.011 (0.939, 1.082)** | **0.552** | **1.18E-111** | 0.474 |
|  | BMFF Diaphysis | Both | **0.818 (0.674, 0.962)** | **0.169** | **1.69E-26** | 0.534 |
| BMFF Total Hip | BMFF Diaphysis | Both | **0.764 (0.669, 0.858)** | **0.281** | **2.48E-48** | **0.001** |
|  |  | Female | **0.65 (0.534, 0.766)** | **0.228** | **8.76E-25** | - |
|  |  | Male | **1.046 (0.857, 1.234)** | **0.331** | **1.03E-22** | - |

### 3.3. Effect of osteopaenia or osteoporosis on BMFF at each site

We next investigated the effect of osteopaenia or osteoporosis on BMFF at each site. As shown in Figure 5, osteopaenic or osteoporotic females had higher BMFF than control females at each site analysed. In males, osteopaenia was associated with significantly increased BMFF at the total hip and femoral diaphysis, and total hip BMFF was also greater osteoporotic vs control males (Fig. 5). However, unlike in females, BMFF at the spine or femoral head did not differ between normal, osteopaenic and osteoporotic males, while diaphyseal BMFF also did not differ between osteoporotic and normal males (Fig. 5).

**Figure 5.**
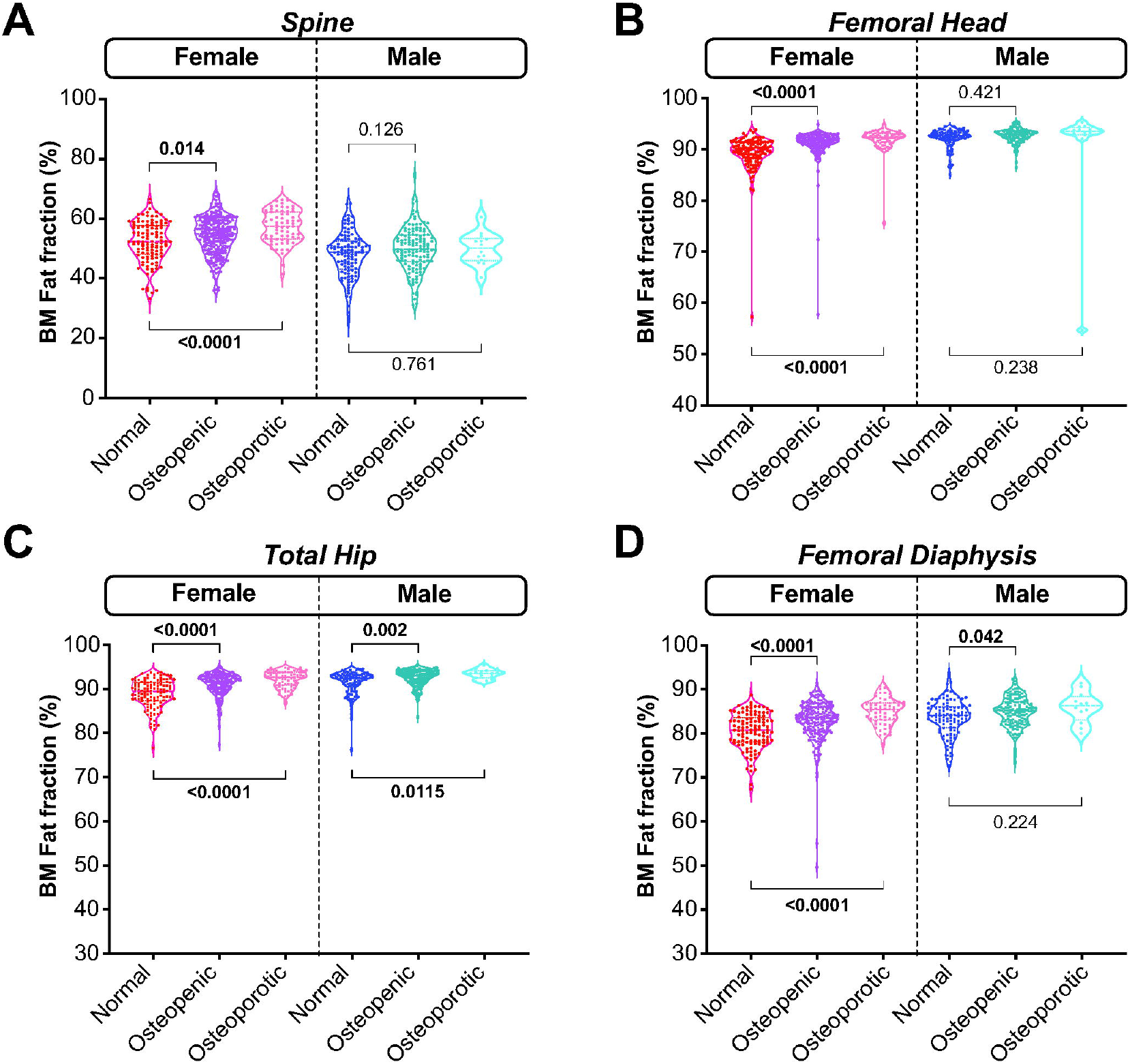
Osteopaenia or osteoporosis influence BMFF in a sex- and region-specific manner. BMFF for control, osteopaenic and osteoporotic subjects was assessed at each skeletal region. Data are shown as violin plots overlaid with individual data points. Within each sex, significant differences between control and osteopaenic or osteoporotic subjects were assessed by one-way ANOVA (for normally distributed data: A) or the Kruskal-Wallis test (for non-normally distributed data: B-D). *P* values for each comparison are shown on each graph.

### 3.4. Univariable associations between BMD, BMFF and other traits

The lack of increased BMFF in osteoporotic males was unexpected and may result from the low numbers in this group (Table 1). Thus, we next used univariable regression to determine if BMFF shows the expected inverse association with BMD at each site, regardless of osteoporotic status. We also investigated which other variables are associated with BMD at each site. As shown in Supplemental Table 1, BMD and BMFF were inversely associated at the spine and this relationship did not differ between the sexes. A similar relationship existed between spine BMD and legs fat %. In contrast, spine BMD was positively associated with visceral adipose tissue (VAT) mass, android fat %, trunk fat % and BMI, with the latter relationship being stronger in males than in females. There was no significant relationship between spine BMD and age, total fat % or gynoid fat %; however, females showed a trend for lower spine BMD with increasing age.

Univariable regression analyses for BMD at the femoral neck, total hip and femoral shaft are presented in Supplemental Tables 2, 3 and 4, respectively. For femoral neck BMD we detected robust inverse associations with BMFF at the femoral head, total hip and spine; the latter relationship was assessed to determine if spinal BMFF is a useful predictor of BMD at the femoral neck, given the clinical significance of fractures at this site. Notably, the relationship with femoral head BMFF showed strong sexual dimorphism, occurring robustly in females while being absent in males. Femoral neck BMD also showed an inverse relationship also with legs fat % and a positive association with BMI; however, no significant associations occurred for the other explanatory variables tested (Supplemental Table 2).

Similar relationships occurred for total hip BMD, including sex differences in the association with femoral head BMFF; an inverse association with legs fat %; and a positive association with BMI (Supplemental Table 3). Unlike for femoral neck BMD, total hip BMD also showed a positive association with VAT mass.

As for these other sites, femoral shaft BMD was inversely associated with BMFF at the femoral diaphysis while being positively associated with BMI. Weaker negative and positive associations were noted for legs fat % and VAT mass, respectively, and none of these relationships differed between the sexes (Supplemental Table 4).

### 3.5. Univariable associations between BMFF and age, BMI or adiposity traits

In addition to BMD, factors including age, BMI and peripheral adiposity have been associated with altered BMFF. Thus, an important question is whether such other factors confound the relationships between BMFF and BMD. To address this, we first used univariable linear regression to identify other variables significantly associated with BMFF at each site, thereby identifying factors associated with BMFF and/or BMD. The results are presented in Supplemental Table 5.

We found that spinal BMFF was positively associated with age, VAT mass, total fat %, android fat %, gynoid fat % and trunk fat % in males and females, with no sex differences in these relationships. In contrast, spinal BMFF showed a positive association with legs fat % in males only (Supplemental Table 5).

Fewer variables were associated with BMFF at the femoral head or total hip. The former showed a positive relationship only with age, and in females only, while the latter was negatively associated only with BMI across both sexes. However, no other variables were associated with BMFF at these two sites (Supplemental Table 5). In contrast, diaphyseal BMFF was associated with several of the variables assessed, often in a sexually dimorphic manner. Thus, across both sexes, diaphyseal BMFF was inversely associated with VAT mass, while inverse associations with total fat %, android fat % and trunk fat % showed significant sex differences, occurring in females but not in males. In contrast, in males, but not females, diaphyseal BMFF was positively associated with legs fat % (Supplemental Table 5).

### 3.6. The inverse association between BMFF and BMD at each site persists after controlling for relevant covariables

Based on the univariable associations identified in Supplemental Tables 1-5, we next constructed multivariable models to estimate the true relationship between BMFF and BMD at each site. Table 4 shows the results for BMD spine as the dependent variable. Here, the best predictive model was obtained when including BMFF Spine, sex, BMI, Legs fat %, VAT mass and Android fat % as covariables (Model 4.6). Notably, the inverse association between spinal BMFF and spinal BMD persisted even when accounting for these other covariables. Moreover, inclusion of leg fat, VAT mass and android fat weakened the size of the sex effect, suggesting that increased spinal BMD in males is explained, at least in part, by their lower amount of leg fat and greater VAT mass and android fat.

**Table 4.** Multivariable regression analyses for spine BMD. Multivariable regression was done using BMD spine as the dependent variable; explanatory variables were selected based on those showing significant univariable association with BMD spine and/or BMFF at the relevant sites, as shown in Supplemental Tables 1-5. For each model the adjusted R^2^ (Adj. R^2^) and Akaike Information Criterion (AIC) are shown, along with multivariable beta coefficients (with lower and upper 95% Cis) for each variable. *P* values are indicated by * (*P*<0.05), ** (*P*<0.01) or *** (*P*<0.001), with significant associations highlighted in bold

|  | *Covariable* |  |  |  |  |  |  |  |
| --- | --- | --- | --- | --- | --- | --- | --- | --- |
|  | Adj. R<sup>2</sup> | AIC | BMFF Spine | Sex (M) | BMI | Legs fat % | VAT mass (kg) | Android fat % |
| Model 4.1 | 0.39 | -893.1 | **-0.004 (-0.006 to -0.003)\*\*\*** | **0.177 (0.156 to 0.198)\*\*\*** | - | - | - | - |
| Model 4.2 | 0.43 | -941.9 | **-0.005 (-0.006, -0.003)\*\*\*** | **0.158 (0.137, 0.179)\*\*\*** | **0.023 (0.017, 0.030)\*\*\*** | - | - | - |
| Model 4.3 | 0.46 | -975.3 | **-0.004 (-0.006, -0.003)\*\*\*** | **0.061 (0.023, 0.098)\*\*** | **0.029 (0.023, 0.036)\*\*\*** | **-0.006 (-0.008, -0.004)\*\*\*** | - | - |
| Model 4.4 | 0.47 | -990.7 | **-0.005 (-0.006, -0.004)\*\*\*** | 0.037 (-0.002, 0.075) | **0.022 (0.015, 0.029)\*\*\*** | **-0.006 (-0.008, -0.004)\*\*\*** | **0.064 (0.033, 0.095)\*\*\*** | - |
| Model 4.5 | 0.47 | -990.1 | **-0.005 (-0.006, -0.004)\*\*\*** | **0.058 (0.020, 0.095)\*\*** | **0.021 (0.014, 0.029)\*\*\*** | **-0.007 (-0.009, -0.005)\*\*\*** | - | **0.003 (0.001, 0.004)\*\*\*** |
| Model 4.6 | 0.47 | -990.9 | **-0.005 (-0.007, -0.004)\*\*\*** | **0.043 (0.004, 0.083)\*** | **0.021 (0.013, 0.028)\*\*\*** | **-0.007 (-0.009, -0.004)\*\*\*** | 0.041 (-0.003, 0.085) | 0.001 (-0.000, 0.003) |

Table 5 shows the results for femoral neck BMD as the dependent variable. Here, separate models were tested for BMFF at the femoral head, total hip or spine as the main explanatory variables; the former was assessed in females only because of the lack of relationship between femoral head BMFF and femoral neck BMD in males (Supplemental Table 2). We found that, in females, the significant inverse association between BMFF femoral head and femoral neck BMD persisted when accounting for BMI and legs fat % (Model 5.3). Similarly, across both sexes, total hip or spine BMFF retained their inverse relationships with femoral neck BMD even after accounting for sex, BMI and legs fat % (Models 5.6 and 5.11). The best model for BMFF total hip also included Android fat % and Trunk fat % (Model 5.8). Notably, male sex was no longer associated with increased femoral neck BMD when controlling for BMFF spine, BMI and legs fat % (Model 5.11), suggesting that males have greater BMD at the femoral neck because they tend to have lower spinal BMFF, lower % leg fat and higher BMI than females.

**Table 5.**
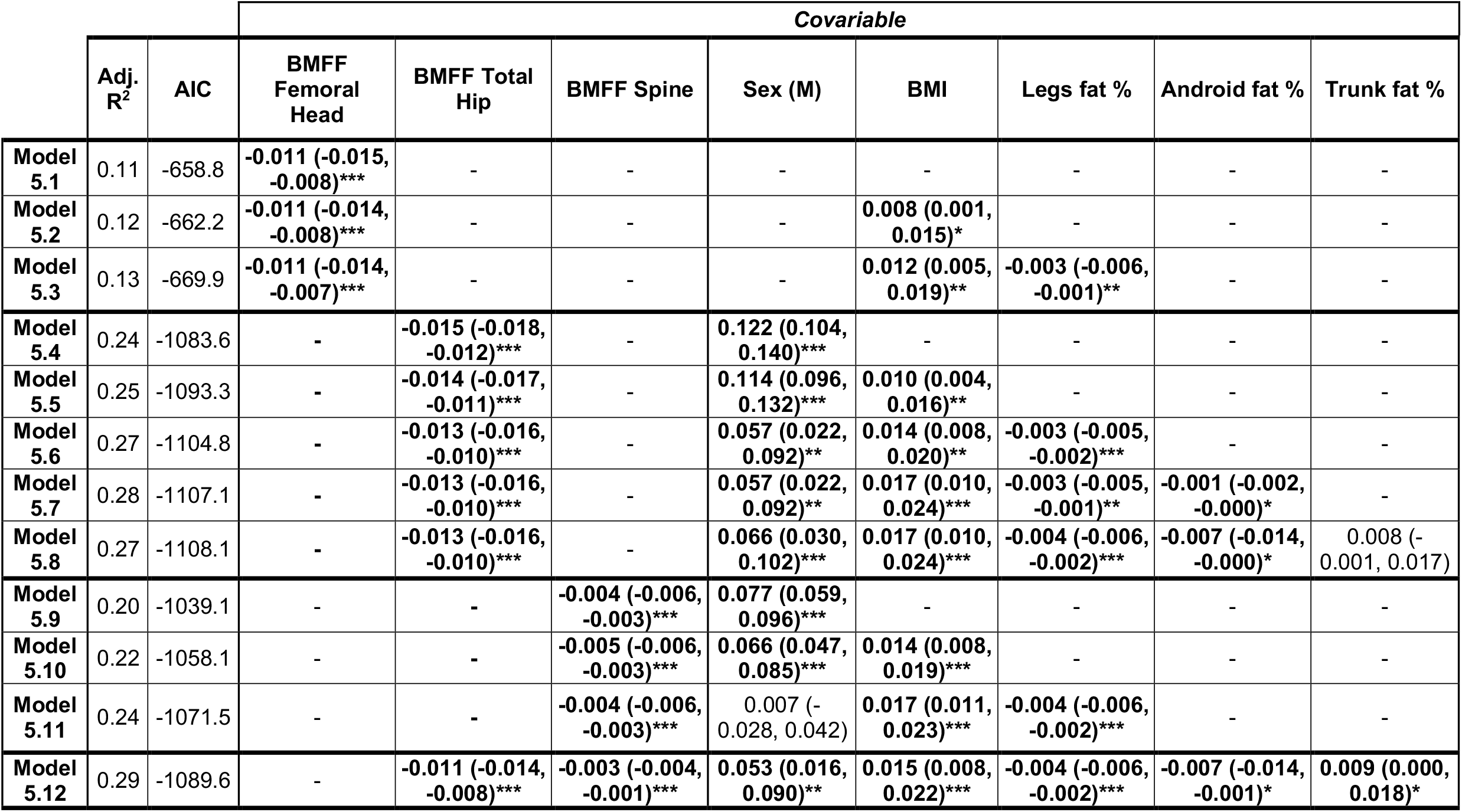
Multivariable regression analyses for femoral neck BMD. Multivariable regression was done using femoral neck BMD as the dependent variable, with BMFF at the femoral head, total hip and spine chosen as the primary explanatory variables. Other explanatory covariables were selected, models constructed, and data presented as described for Table 4. Models with femoral head BMFF (5.1, 5.2 and 5.3) were tested in females only because univariable analysis showed that this is not associated total hip BMD in males (Supplemental Table 2).

Given that spine BMFF is positively associated with total hip BMFF (Table 3), we postulated that the inverse relationship between spine BMFF and femoral neck BMD may occur because spine BMFF is a surrogate for total hip BMFF. However, the inverse relationship between spine BMFF and femoral neck BMD persisted even when accounting for BMFF at the total hip (Model 5.12), demonstrating that these explanatory variables are acting at least partly independently of each other.

Multivariable regression for total hip BMD is presented in Table 6. The best predictive model included BMFF total hip, sex, BMI and legs fat % as the covariables (Model 6.3); inclusion of VAT mass (Model 6.4) did not further improve the model, despite VAT mass showing a significant univariable association with total hip BMD (Supplemental Table 3). Notably, the inverse relationship between total hip BMD and BMFF persisted even when accounting for sex, BMI and legs fat %, confirming total hip BMFF as an independent predictor of BMD at this site.

**Table 6.** Multivariable regression analyses for total hip BMD. Multivariable regression was done using total hip BMD as the dependent variable, with BMFF at the total hip as the primary explanatory variable. Other explanatory covariables were selected, models constructed, and data presented as described for Table 4.

|  | Adj.<br>R<sup>2</sup> | AIC | Covariable |  |  |  |  |
| --- | --- | --- | --- | --- | --- | --- | --- |
|  |  |  | BMFF<br>Total Hip | Sex (M) | BMI | Legs fat<br>% | VAT<br>mass (kg) |
| **Model 6.1** | 0.34 | -997.9 | **-0.017 (-0.020, -0.014)\*\*\*** | **0.170 (0.152, 0.189)\*\*\*** | - | - | - |
| **Model 6.2** | 0.37 | -1029.2 | **-0.016 (-0.019, -0.013)\*\*\*** | **0.156 (0.137, 0.175)\*\*\*** | **0.018 (0.012, 0.023)\*\*\*** | - | - |
| **Model 6.3** | 0.39 | -1056.6 | **-0.015 (-0.018, -0.012)\*\*\*** | **0.069 (0.033, 0.105)\*\*\*** | **0.023 (0.017, 0.029)\*\*\*** | **-0.005 (-0.007, -0.003)\*\*\*** | - |
| **Model 6.4** | 0.39 | -1051.3 | **-0.015 (-0.018, -0.012)\*\*\*** | **0.074 (0.037, 0.111)\*\*\*** | **0.025 (0.018, 0.032)\*\*\*** | **-0.005 (-0.007, -0.003)\*\*\*** | **-0.016 (-0.045, 0.012)** |

Finally, Table 7 shows the results of multivariable regression for femoral shaft BMD. Here, the best predictive model included diaphyseal BMFF, sex, BMI, legs fat % and android fat % (Model 7.5), although a similarly accurate model was obtained when VAT mass and trunk fat % were also included (Model 7.7). As for the other BMFF-BMD relationships, BMFF at the diaphysis retained its significant inverse association with femoral shaft BMD even when these other covariables were accounted for. Moreover, males no longer had significant increases in femoral shaft BMD when controlling for BMFF diaphysis, BMI and legs fat % (Model 7.3-7.7). This suggests that males may have greater femoral shaft BMD because they have a higher BMI and lower % leg fat than females.

**Table 7.** Multivariable regression analyses for femoral shaft BMD. Multivariable regression was done using femoral shaft BMD as the dependent variable; explanatory covariables were selected, models constructed, and data presented as described for Table 4.

|  | Covariable |  |  |  |  |  |  |  |  |
| --- | --- | --- | --- | --- | --- | --- | --- | --- | --- |
|  | Adj. R<sup>2</sup> | AIC | BMFF Diaphysis | Sex (M) | BMI | Legs fat % | VAT mass (kg) | Android fat % | Trunk fat % |
| Model 7.1 | 0.28 | -693.2 | -0.014 (-0.016, -0.011)*** | 0.164 (0.141, 0.187)*** | - | - | - | - | - |
| Model 7.2 | 0.30 | -711.7 | -0.013 (-0.016, -0.011)*** | 0.151 (0.127, 0.174)*** | 0.017 (0.010, 0.025)*** | - | - | - | - |
| Model 7.3 | 0.33 | -743.1 | -0.013 (-0.016, -0.011)*** | 0.035 (-0.010, 0.080) | 0.025 (0.017, 0.032)*** | -0.007 (-0.010, -0.005)*** | - | - | - |
| Model 7.4 | 0.33 | -739.5 | -0.013 (-0.016, -0.011)*** | 0.045 (-0.002, 0.091) | 0.028 (0.019, 0.037)*** | -0.007 (-0.010, -0.005)*** | -0.029 (-0.065, 0.007) | - | - |
| Model 7.5 | 0.34 | -747.1 | -0.014 (-0.016, -0.011)*** | 0.038 (-0.007, 0.083) | 0.030 (0.021, 0.039)*** | -0.006 (-0.009, -0.004)*** | - | -0.002 (-0.004, -0.000)* | - |
| Model 7.6 | 0.34 | -745.3 | -0.013 (-0.016, -0.011)*** | 0.035 (-0.010, 0.080) | 0.030 (0.021, 0.039)*** | -0.006 (-0.009, -0.004)*** | - | - | -0.002 (-0.004, -0.000)* |
| Model 7.7 | 0.34 | -743.4 | -0.014 (-0.016, -0.011)*** | 0.047 (-0.002, 0.096) | 0.030 (0.020, 0.039)*** | -0.008 (-0.011, -0.005)*** | 0.007 (-0.044, 0.059) | -0.011 (-0.020, -0.002)* | 0.012 (0.000, 0.024)* |

## 4. DISCUSSION

Herein, we have developed a new deep learning method for analysis of BM adiposity using Dixon MRI data from the UKBB. This is the first study to establish deep learning for BM segmentation at multiple sites, and the first do so, for any skeletal site, in the UKBB imaging study. Our models yield BMFF measurements that are consistent with previous observations, including sex differences in spinal BMFF and inverse associations with BMD. This demonstrates the ability of our models to generate accurate, reliable BMFF measurements from the UKBB MRI data. We further reveal new site- and sex-specific associations that have not been reported previously, highlighting the potential of our methods to uncover new pathophysiological functions of BMAT.

### 4.1. Deep learning for large-scale BM analysis

Several other recent studies have developed deep learning for automated BM segmentation from MRI data. For example, von Brandis *et al* assessed the feasibility of deep learning for segmenting BM from T2-weighted Dixon water-only images, focusing on the knee region (20); however, the best median dice score of their model was only 0.68, far below that obtained by our models (Table 2). Better accuracy was achieved by Zhou *et* al, who established a deep learning model for segmenting lumbar vertebrae from Dixon MRI data (18). They trained their model using manual segmentations of 165 vertebrae from 31 subjects, with the model then tested on a validation set of 24 subjects. They achieved an average dice score of 0.849, below the accuracy of our vertebral ROI-Attention-U-Net (Table 2). More recently, Zhao *et al* used deep learning for segmenting lumbar vertebrae from modified Dixon MRI data, using a training set of 142 subjects and a validation set of 64 subjects (19). Their model achieved a mean dice score of 0.912, the same as that obtained by our vertebral ROI-Attention-U-Net (Table 2). Thus, among deep learning models for segmenting vertebral BM, our model achieves an accuracy that is similar or greater than that obtained by others.

Notably, our study is the first to develop deep learning for BM segmentation at the femoral head, total hip and femoral diaphysis. This is important because the properties of BMAT vary according to skeletal location (1, 7, 8). Thus, to fully understand the health implications of BMAT and its potential utility as a clinical biomarker, it is critical to assess BMFF at other sites. Indeed, as discussed below, we found that the associations between BMFF, age, BMD, BMI and peripheral adiposity differ according to the BM region assessed, underscoring the importance of assessing BMFF across multiple sites. Finally, our model includes dedicated error-checking steps to remove inaccurate segmentation outputs, which is essential for reliable analysis of large-scale MRI data.

### 4.2. New ROI attention U-Net model

Another advance of the present study is our development of a new ROI attention U-Net model that allows accurate segmentation of small VOIs from large volumetric data. The traditional 3D U-Net has a fixed receptive field that is dependent on the size of convolutional kernels and network depth. To achieve state-of-the-art performance, the network architecture needs to be carefully designed to fit the sizes of the segmented objects and image resolution. As a result, in this study the traditional 3D U-Net generates highly accurate results for vertebrae and femoral head (Table 2), regions in which the segmented objects are relatively large. However, this traditional U-Net shows limited discriminative power when dealing with smaller structures such as the femoral diaphysis, where only a few pixels on each axial slice are annotated as foreground. On the contrary, our new ROI attention U-Net model can adaptively encode the local and global contextual information with its adjustive-attention mechanism. As shown in Table 2, it increased segmentation accuracy of the femoral diaphysis by over 25% and also slightly improved accuracy for the total hip region. Alongside these improvements, for the femoral head and vertebrae the ROI attention U-Net performs similarly to the carefully designed traditional 3D U-Net (Table 2). Thus, our new ROI attention model advances the state of the art by achieving accurate segmentation of both larger and smaller objects.

### 4.3. Association between BMFF and pathophysiological characteristics – confirmation of previous studies and new findings

The key aim of our study was to develop and validate deep learning models for automated BM segmentation of UKBB Dixon MRI data. Our group of 729 subjects is the largest cohort yet to undergo measurement of spinal BMFF, and by far the largest to include assessment of BMFF at any femoral site (12). Consistent with previous reports, we find that spinal BMFF is lower than femoral BMFF (Fig. 4) (1, 12, 31); is greater in females than in males (Fig. 4) (29, 30); increases with age (Supplemental Table 5) (12, 29, 30, 32); is elevated in osteopaenia or osteoporosis, at least in females (Fig. 5) (1, 6, 12); exhibits a robust, inverse association with spinal BMD (Table 4) (1, 6, 12); and is positively associated with visceral adiposity (Supplemental Table 5) (32, 33).

Our results for femoral BMFF are also consistent with previous studies. For example, in a cohort of aged females, Griffith *et al* found that BMFF in the femoral head, neck and diaphysis is increased in osteoporosis and inversely associated with BMD at each site (34). We confirm these findings (Fig. 5, Tables 5-7) and further reveal that femoral head BMFF is not associated with BMD at the femoral neck or total hip in males (Supplemental Tables 2-3). We also show that diaphyseal BMFF is typically inversely associated with peripheral adiposity in females but not in males, while BMFF at the femoral head or total hip is not associated with these peripheral adiposity traits (Supplemental Table 5); these observations confirm and extend those of a previous smaller-scale study (35). The reasons for these variable site- and sex-dependent relationships between BMFF and peripheral adiposity remain to be determined; however, one possibility is that they reflect preferences for the partitioning of lipid storage between different adipose depots.

Many of our new findings relate to the fact that most previous MR-based studies of BM adiposity have focussed on vertebrae, with femoral sites being relatively overlooked (12). For example, we show that, across both sexes, BMFF is highest in the femoral head and decreases progressively in the total hip and diaphysis, while BMFF at each femoral site is greater in males than in females (Fig. 4). Unlike in the spine, age is associated with increased femoral head BMFF only in females, and across both sexes shows no relationship with total hip or diaphyseal BMFF (Supplemental Table 5). This could reflect the fact that, compared to the spine, these femoral sites contain a greater proportion of constitutive BMAT, which is less age responsive than the regulated BMAT that predominates in the axial skeleton (7, 8). However, it may be that age-related increases in femoral BMAT occur over a longer timeframe that would only be apparent when BMFF is assessed over a greater age range.

Regarding constitutive vs regulated subtypes, we also find robust positive associations between BMFF at the four different sites analysed (Table 3), similar to the findings of Slade *et al* (31). However, we further reveal that these relationships exhibit sex differences and are strongest between the three femoral regions, with spinal BMFF showing no association with diaphyseal BMFF (Table 3). This may reflect differences in the development and function of regulated vs constitutive BMAT (7, 8).

Together, our present findings confirm those of previous studies while also revealing new knowledge about BMAT’s site- and sex-dependent characteristics. This underscores the ability of our deep learning models to yield reliable BMFF measurements and to identify new insights into the pathophysiology of BMAT.

### 4.4. Limitations

One specific limitation is that our cohort included relatively few osteoporotic males. This restricted our ability, in males, to detect significant effects of osteoporosis on BMFF at each site. Our univariable and multivariable regression analyses were still able to detect significant inverse associations between BMFF and BMD at each site; however, once we have measured BMFF across the full available UKBB cohort it will be informative to reassess the relationship between BMFF and osteoporosis.

A more-general limitation relates to the UKBB MRI protocol. Participants in the UKBB imaging study visited several different imaging centres for acquisition of the MRI scans. Therefore, across these different imaging centres the MRI protocol parameters had to be standardised and harmonised, resulting in both advantages and drawbacks. For example, to simplify the procedure the Dixon sequences were based on only two echo times; however, with only dual-echo sequences, no accurate T2*-correction could be applied and the complexity of the fat spectrum could not be considered in the BMFF mapping (10, 14). As a result, reported BMFF measurements can be affected by T2* decay effects caused by the presence of trabecular bone, which in turn may differ in the water and fat components (9, 10). However, the moderately low flip angle (10°) is acceptable to limit T1-bias, and protocol standardisation compelled all examinations to be performed in similar conditions, with the exact same parameters (9, 36). Consequently, even if the true proton-density fat fraction (PDFF) could not be quantified, a comparable estimate could be obtained through the reported BMFF, which permits group comparison and method cross-validation. Furthermore, dual-echo Dixon-derived BMFF allows the derivation of consistent 3D BMFF measurements across all UKBB MR imaging centres. This is very important for our BMFF validation study, as it allowed us to assess and automate extraction of BMFF maps from multiple skeletal sites, on a 3D mode.

### 4.5. Conclusions

Our new deep learning models allow accurate segmentation of small VOIs from large volumetric MRI data. While we have used these models to analyse small BM regions, they could also be applied for precise, automated, large-scale analysis of other small anatomical structures of interest. The development and validation of our models using UKBB MRI data is hugely significant because, unlike most other MRI datasets, the UKBB also provides extensive genetic and phenotypic data for each subject, including whole-genome sequencing and health records. This linked data allows comprehensive association studies to identify the genetic and pathophysiological factors associated with FF and other MRI-derived measurements. Indeed, Liu *et al* recently demonstrated the power of this approach using deep learning for segmentation of abdominal organs from UKBB MRI data (16). They identified genetic variants and clinical conditions associated with FF and other imaging-derived characteristics for each organ, as well as combinations of characteristics across multiple organs. The deep learning models established in the present study unlock similar possibilities: using these new models, we will next measure BMFF across the full UKBB imaging cohort, which will eventually include 100,000 subjects. This will allow us to identify the genetic, physiological and clinical conditions associated with altered BMFF at each site. Such knowledge will help to elucidate the mechanisms that influence BM adiposity and reveal, to an unprecedented extent, how BMAT impacts human health and disease.

## Data Availability

All data for fat fraction (FF) and segmentation volumes will be uploaded to the UK Biobank. Code for the deep learning models will be made available via GitHub. Code for regression analyses will be made available via DataShare (https://datashare.ed.ac.uk). Until these data are publicly available, the authors will agree to all reasonable requests for code and data sharing, in accordance with UK Biobank guidelines.

## ACKNOWLEDGEMENTS

This work was supported by a grant from the Medical Research Council (MR/S010505/1 to W.P.C.). W.P.C. was further supported by a Chancellor’s Fellowship from the University of Edinburgh. C.W. was further supported by the British Heart Foundation (RG/16/10/32375). C.G. and T.M. were supported by the Edinburgh Clinical Research Facility and NHS Lothian R&D.

We are grateful to Dominic Job (Edinburgh Imaging, University of Edinburgh) for support with IT infrastructure, and Jimmy Bell, Louise Thomas and Brandon Whitcher (University of Westminster) for helpful discussions and advice regarding working with UKBB MRI data.

## AUTHOR CONTRIBUTIONS (based on <u>CRediT taxononmy</u>)

***Conceptualisation***, W.P.C.; ***Data curation***, D.M.M., C.W., G.P. and W.P.C.; ***Formal Analysis***, D.M.M., C.W., G.P., C.D.G. and W.P.C.; ***Funding Acquisition***,, S.I.K.S., T.M. and W.P.C.; ***Investigation***, D.M.M., C.W., G.P. and W.P.C.; ***Methodology***, D.M.M., C.W., G.P., C.D.G., S.B., J.P., S.I.K.S., T.M. and W.P.C.; **Project administration**, S.I.K.S., T.M. and W.P.C.; ***Resources***, S.I.K.S., T.M. and W.P.C.; ***Software***, D.M.M., C.W., G.P.; ***Supervision***, S.I.K.S., T.M. and W.P.C.; ***Visualisation***, D.M.M., C.W., C.D.G. and W.P.C.; ***Writing – Original Draft***, D.M.M., C.W., S.B., J.P. and W.P.C..; ***Writing – Review & Editing***, D.M.M., C.W., G.P., C.D.G., S.B., J.P., S.I.K.S., T.M. and W.P.C.

