## Supplemental Tables 1-5 for "A novel deep learning method for large-scale analysis of bone marrow adiposity using UK Biobank Dixon MRI data"

Morris et al.

### SUPPLEMENTAL DIGITAL CONTENT

The Supplemental Digital Content includes five tables, as follows:

**Supplemental Table 1** – Univariable associations between spine BMD and BMFF, Age, BMI or adiposity traits.

**Supplemental Table 2** – Univariable associations between femoral head BMD and BMFF, Age, BMI or adiposity traits.

**Supplemental Table 3** – Univariable associations between total hip BMD and BMFF, Age, BMI or adiposity traits.

**Supplemental Table 4** – Univariable associations between femoral shaft BMD and BMFF, Age, BMI or adiposity traits.

**Supplemental Table 5** – Univariable associations between BMFF at each region and Age, BMI or adiposity traits.

| Dependent | Explanatory | Sex | β (95% CIs) | Adj. $R^2$ | *P* (Exp) | *P* (Exp*Sex) |
|---|---|---|---|---|---|---|
| BMD Spine | BMFF Spine | Both | **-0.004 (-0.006, -0.003)** | **0.390** | **1.09E-09** | 0.431 |
| | Age | Both | 0 (-0.004, 0.003) | 0.352 | 0.930 | **0.021** |
| | | F | -0.003 (-0.007, 0.001) | 0.003 | 0.116 | - |
| | | M | 0.005 (-0.001, 0.012) | 0.006 | 0.108 | - |
| | BMI | Both | **0.023 (0.017, 0.029)** | **0.395** | **1.606E-12** | **7.827E-05** |
| | | F | **0.015 (0.008, 0.022)** | **0.036** | **3.276E-05** | - |
| | | M | **0.043 (0.03, 0.055)** | **0.137** | **1.254E-10** | - |
| | VAT mass (kg) | Both | **0.081 (0.054, 0.109)** | **0.380** | **1.01E-08** | 0.840 |
| | Total fat % | Both | 0.002 (0, 0.004) | 0.354 | 1.19E-01 | 0.438 |
| | Android fat % | Both | **0.002 (0.001, 0.004)** | **0.369** | **9.25E-06** | 0.904 |
| | Gynoid fat % | Both | -0.002 (-0.004, 0) | 0.354 | 0.096 | 0.208 |
| | Trunk fat % | Both | **0.003 (0.001, 0.004)** | **0.365** | **8.84E-05** | 0.716 |
| | Legs fat % | Both | **-0.004 (-0.006, -0.001)** | **0.362** | **6.78E-04** | 0.276 |

**Supplemental Table 1 – Univariable associations between spine BMD and BMFF, Age, BMI or adiposity traits.** To test if the explanatory-dependent relationship differs between males and F, a linear model was first analysed across both sexes, with sex included as an interacting variable. Beta coefficients are shown (with lower and upper 95% CIs in brackets), followed by the adjusted $R^2$ (Adj. $R^2$) and unadjusted *P* value for each explanatory variable (*P* Exp). *P* values were also calculated for the Explanatory*Sex interaction (*P* Exp*Sex); if significant, additional linear models were analysed in females (F) and males (M) separately. Because 13 correlations were assessed, the Bonferroni-adjusted alpha level for *P* (Exp) is 0.05/13 = 0.0038. Significant explanatory-dependent relationships are highlighted in bold.

| Dependent | Explanatory | Sex | β (95% CIs) | Adj. $R^2$ | *P* (Exp) | *P* (Exp*Sex) |
|---|---|---|---|---|---|---|
| BMD Femoral Neck | BMFF Femoral Head | Both | **-0.008 (-0.011, -0.005)** | **0.178** | **4.73E-08** | **2.100E-04** |
| | | F | **-0.011 (-0.015, -0.008)** | **0.107** | **8.834E-12** | **-** |
| | | M | 0 (-0.005, 0.005) | -0.004 | 0.954 | - |
| | BMFF Total Hip | Both | **-0.015 (-0.018, -0.012)** | **0.243** | **3.498E-20** | 0.404 |
| | BMFF Spine | Both | **-0.004 (-0.006, -0.003)** | **0.201** | **6.728E-12** | 0.886 |
| | Age | Both | -0.002 (-0.005, 0.001) | 0.142 | 0.258 | 0.809 |
| | BMI | Both | **0.014 (0.008, 0.02)** | **0.167** | **2.173E-06** | 0.070 |
| | VAT mass (kg) | Both | 0.021 (-0.005, 0.046) | 0.143 | 0.113 | 0.572 |
| | Total fat % | Both | -0.001 (-0.002, 0.001) | 0.141 | 0.438 | 0.438 |
| | Android fat % | Both | 0 (-0.001, 0.001) | 0.140 | 0.760 | 0.381 |
| | Gynoid fat % | Both | -0.002 (-0.004, 0) | 0.144 | 0.087 | 0.693 |
| | Trunk fat % | Both | 0 (-0.001, 0.001) | 0.140 | 0.768 | 0.553 |
| | Legs fat % | Both | **-0.003 (-0.004, -0.001)** | **0.149** | **0.008** | 0.323 |

**Supplemental Table 2 – Univariable associations between femoral head BMD and BMFF, Age, BMI or adiposity traits.** Linear models were established as described for Supplemental Table 1. Because 13 correlations were assessed, the Bonferroni-adjusted alpha level for *P* (Exp) is 0.05/13 = 0.0038. Significant explanatory-dependent relationships are highlighted in bold.

| Dependent | Explanatory | Sex | β (95% CIs) | Adj. $R^2$ | *P* (Exp) | *P* (Exp*Sex) |
|---|---|---|---|---|---|---|
| BMD Total Hip | BMFF Femoral Head | Both | **-0.008 (-0.011, -0.005)** | **0.268** | **7.06E-07** | **0.007** |
| | | F | **-0.01 (-0.014, -0.007)** | **0.079** | **4.564E-09** | - |
| | | M | -0.002 (-0.007, 0.004) | -0.003 | 0.575 | - |
| | BMFF Total Hip | Both | **-0.017 (-0.02, -0.014)** | **0.339** | **2.408E-23** | 0.078 |
| | Age | Both | 0.002 (-0.005, 0.001) | 0.234 | 0.226 | 0.770 |
| | BMI | Both | **0.022 (0.016, 0.028)** | **0.282** | **3.044E-12** | 0.111 |
| | VAT mass (kg) | Both | **0.044 (0.017, 0.071)** | **0.242** | **0.002** | 0.979 |
| | Total fat % | Both | 0 (-0.002, 0.002) | 0.232 | 0.929 | 0.175 |
| | Android fat % | Both | 0.001 (0, 0.002) | 0.235 | 0.091 | 0.225 |
| | Gynoid fat % | Both | -0.002 (-0.004, 0) | 0.236 | 0.043 | 0.248 |
| | Trunk fat % | Both | 0.001 (0, 0.002) | 0.235 | 0.106 | 0.240 |
| | Legs fat % | Both | **-0.003 (-0.005, -0.001)** | **0.244** | **6.93E-04** | 0.115 |

**Supplemental Table 3 – Univariable associations between total hip BMD and BMFF, Age, BMI or adiposity traits.** Linear models were established as described for Supplemental Table 1. Because 12 correlations were assessed, the Bonferroni-adjusted alpha level for *P* (Exp) is 0.05/12 = 0.0042. Significant explanatory-dependent relationships are highlighted in bold.

| Dependent | Explanatory | Sex | β (95% CIs) | Adj. $R^2$ | *P* (Exp) | *P* (Exp*Sex) |
|---|---|---|---|---|---|---|
| BMD Femoral shaft | BMFF Femoral Diaphysis | Both | **-0.014 (-0.016, -0.011)** | **0.277** | **2.59E-23** | 0.640 |
| | Age | Both | -0.004 (-0.008, 0.001) | 0.152 | 0.086 | 0.439 |
| | BMI | Both | **0.024 (0.016, 0.032)** | **0.191** | **1.319E-09** | 0.337 |
| | VAT mass (kg) | Both | **0.046 (0.012, 0.08)** | **0.156** | **0.008** | 0.983 |
| | Total fat % | Both | 0 (-0.003, 0.002) | 0.149 | 0.870 | 0.419 |
| | Android fat % | Both | 0.001 (0, 0.002) | 0.151 | 0.149 | 0.355 |
| | Gynoid fat % | Both | -0.002 (-0.005, 0) | 0.153 | 0.062 | 0.579 |
| | Trunk fat % | Both | 0.001 (0, 0.003) | 0.151 | 0.153 | 0.409 |
| | Legs fat % | Both | **-0.004 (-0.007, -0.002)** | **0.161** | **0.001** | 0.427 |

**Supplemental Table 4 – Univariable associations between femoral shaft BMD and BMFF, Age, BMI or adiposity traits.** Linear models were established as described for Supplemental Table 1. Because 9 correlations were assessed, the Bonferroni-adjusted alpha level for *P* (Exp) is 0.05/9 = 0.0056. Significant explanatory-dependent relationships are highlighted in bold.

| Dependent | Explanatory | Sex | β (95% CIs) | Adj. $R^2$ | *P* (Exp) | *P* (Exp*Sex) |
|---|---|---|---|---|---|---|
| BMFF Spine | **Age** | Both | **0.269 (0.08, 0.459)** | **0.127** | **0.005** | 0.542 |
| | BMI | Both | 0.219 (-0.138, 0.576) | 0.119 | 0.229 | 0.425 |
| | **VAT mass (kg)** | Both | **5.331 (3.833, 6.829)** | **0.177** | **6.640E-12** | 0.549 |
| | **Total fat %** | Both | **0.372 (0.267, 0.476)** | **0.176** | **6.492E-12** | 0.075 |
| | **Android fat %** | Both | **0.226 (0.167, 0.285)** | **0.185** | **1.14E-13** | 0.157 |
| | **Gynoid fat %** | Both | **0.241 (0.129, 0.352)** | **0.140** | **2.53E-05** | 0.078 |
| | **Trunk fat %** | Both | **0.287 (0.214, 0.36)** | **0.188** | **3.73E-14** | 0.214 |
| | Legs fat % | Both | 0.141 (0.03, 0.252) | 0.125 | 0.013 | **0.011** |
| | | F | 0.061 (-0.06, 0.183) | 0.000 | 0.323 | - |
| | | M | **0.4 (0.159, 0.641)** | **0.037** | **0.001** | - |
| BMFF Femoral Head | **Age** | Both | 0.097 (0.007, 0.187) | 0.058 | 0.036 | **0.045** |
| | | F | **0.163 (0.048, 0.278)** | **0.016** | **0.005** | - |
| | | M | 0.031 (-0.175, 0.114) | 0.004 | 0.678 | - |
| | BMI | Both | -0.1 (-0.272, 0.071) | 0.054 | 0.251 | 0.291 |
| | VAT mass (kg) | Both | 0.034 (-0.715, 0.782) | 0.052 | 0.930 | 0.876 |
| | Total fat % | Both | 0.043 (-0.01, 0.096) | 0.056 | 0.110 | 0.906 |
| | Android fat % | Both | 0.009 (-0.021, 0.039) | 0.052 | 0.544 | 0.369 |
| | Gynoid fat % | Both | 0.072 (0.016, 0.127) | 0.061 | 0.011 | 0.474 |
| | Trunk fat % | Both | 0.016 (-0.021, 0.053) | 0.053 | 0.402 | 0.516 |
| | Legs fat % | Both | 0.07 (0.016, 0.123) | 0.062 | 0.011 | 0.583 |
| BMFF Total Hip | Age | Both | 0.074 (0, 0.149) | 0.072 | 0.050 | 0.283 |
| | **BMI** | Both | **-0.235 (-0.373, -0.097)** | **0.082** | **8.83E-04** | 0.873 |
| | VAT mass (kg) | Both | -0.665 (-1.265, -0.065) | 0.074 | 0.030 | 0.871 |
| | Total fat % | Both | 0.006 (-0.036, 0.048) | 0.067 | 0.775 | 0.592 |
| | Android fat % | Both | -0.008 (-0.032, 0.015) | 0.068 | 0.492 | 0.996 |
| | Gynoid fat % | Both | 0.032 (-0.012, 0.076) | 0.070 | 0.155 | 0.443 |
| | Trunk fat % | Both | -0.006 (-0.035, 0.023) | 0.067 | 0.686 | 0.719 |
| | Legs fat % | Both | 0.036 (-0.007, 0.08) | 0.071 | 0.103 | 0.847 |
| BMFF Femoral Diaphysis | Age | Both | 0.109 (-0.01, 0.227) | 0.047 | 0.072 | 0.642 |
| | BMI | Both | 0.023 (-0.425, 0.019) | 0.047 | 0.073 | 0.521 |
| | **VAT mass (kg)** | Both | **-1.289 (-2.249, -0.329)** | **0.053** | **0.009** | 0.146 |
| | **Total fat %** | Both | **-0.1 (-0.167, -0.033)** | **0.054** | **0.004** | **0.005** |
| | | F | **-0.164 (-0.25, -0.079)** | **0.031** | **1.81E-04** | - |
| | | M | 0.04 (-0.066, 0.146) | -0.002 | 0.456 | - |
| | **Android fat %** | Both | **-0.075 (-0.112, -0.037)** | **0.064** | **9.415E-05** | **0.013** |
| | | F | **-0.107 (-0.155, -0.059)** | **0.043** | **1.403E-05** | - |
| | | M | -0.007 (-0.066, 0.052) | -0.004 | 0.823 | - |
| | Gynoid fat % | Both | 0.001 (-0.07, 0.072) | 0.042 | 0.983 | **0.022** |
| | | F | -0.055 (-0.146, 0.037) | 0.001 | 0.240 | - |
| | | M | 0.125 (0.013, 0.236) | 0.015 | 0.029 | - |
| | **Trunk fat %** | Both | **-0.09 (-0.137, -0.044)** | **0.063** | **1.606E-04** | **0.013** |
| | | F | **-0.131 (-0.19, -0.071)** | **0.041** | **2.105E-05** | - |
| | | M | -0.004 (-0.077, 0.07) | -0.004 | 0.920 | - |
| | **Legs fat %** | Both | 0.009 (-0.062, 0.079) | 0.042 | 0.809 | **0.004** |
| | | F | -0.048 (-0.134, 0.038) | 0.001 | 0.273 | - |
| | | M | **0.191 (0.066, 0.316)** | **0.032** | **0.003** | - |

**Supplemental Table 5 – Univariable associations between BMFF at each region and Age, BMI or adiposity traits.** Linear models were established as described for

Supplemental Table 1. For each dependent variable, Bonferroni-adjusted alpha levels for *P* (Exp) are as follows: BMFF Spine, 0.005 (10 comparisons); BMFF femoral head, 0.005 (10 comparisons); BMFF total hip, 0.00625 (8 comparisons); BMFF femoral diaphysis, 0.028 (18 comparisons). Significant explanatory-dependent relationships are highlighted in bold.
